# BMI inadequately mediates the relationship between polygenic risk of adiposity and early onset type 2 diabetes in south Asians

**DOI:** 10.1101/2025.06.12.25329480

**Authors:** Binur Orazumbekova, Julia Zöllner, Sam Hodgson, Margherita Bigossi, Miriam Samuel, Genes and Health Research Team, Sarah Finer, Rohini Mathur, Moneeza K Siddiqui

## Abstract

**Aims:** We investigated the relationship between polygenic score (PGS) for BMI and other adiposity PGS with age at type 2 diabetes onset in white European (EUR) and south Asian (SAS) ancestries, and the mediating role of BMI.

**Methods:** In this retrospective study, using polygenic score (PGS) for BMI, clinically measured BMI and age at type 2 diabetes onset, we conducted mediation analysis separately for SAS (n=3,901, Genes & Health) and EUR (n=729, UK Biobank) aged 40 years or older. For SAS, we also used multivariable linear regression with backward selection to identify the best adiposity PGS (waist circumference (WC), waist-to-hip ratio (WHR), visceral adipose tissue (VAT), body fat (BF), trunk fat (TF) and gluteofemoral fat (GFAT), hand-grip strength (HGS)) predicting age at type 2 diabetes onset.

**Results:** A one SD increment in BMI-PGS was associated with earlier type 2 diabetes onset by −0.73 years (95%CI −1.01; −0.45) in SAS and −0.57 years (95%CI −1.05; −0.08) in EUR. BMI fully mediated the PGS effect in EUR (100%) and only partially in SAS (28%). Alongside BMI-PGS, WC-PGS and TF-PGS were good at discriminating measured BMI, WC and WHTR in SAS and were correlated with BMI-PGS. Other best predictors of early onset type 2 diabetes in SAS were WC-PGS, WHR-PGS, BF-PGS and GFAT-PGS, which differed between SAS subgroups and by sex.

**Conclusions:** These findings underscore the importance of incorporating adiposity-related genetics in predicting type 2 diabetes onset among SAS and demonstrate the limitations of using BMI alone to capture associated risk, particularly in diverse populations with typically lower BMI.

**Research in context:** **What is already known about this subject?**

- South Asians develop type 2 diabetes, on average, a decade earlier than white Europeans and at lower BMI levels.
- The well-established association between BMI and type 2 diabetes risk in white Europeans is more complex in south Asians, who have different patterns of adipose tissue distribution that are not well reflected in BMI.
- The contribution of adiposity-related polygenic scores (PGS) to type 2 diabetes onset has not been examined across ancestries.

**What is the key question?**

- What is the association between adiposity PGS and type 2 diabetes onset across ancestries, what how much of this effect is mediated by BMI?

**What are the new findings?**

- Higher BMI-PGS is associated with earlier type 2 diabetes onset in both south Asians and white Europeans; this relationship is fully mediated by BMI in white Europeans but only partially in south Asians.
- Other PGS for central adiposity are significant predictors of early type 2 diabetes onset and central adiposity anthropometrics in south Asians.

**How might this impact on clinical practice in the foreseeable future?**

- These findings highlight the importance of incorporating adiposity-related genetics in predicting type 2 diabetes onset in diverse populations and underscore the need to move beyond BMI when assessing metabolic risk and potentially evaluating the effectiveness of interventions.

## Introduction

Type 2 diabetes is a major public health issue, affecting 589 million people globally [1]. South Asians (SAS), who make up a quarter of the world’s population, bear a disproportionately higher burden of the disease [2]. Almost one-fourth of the global population living with type 2 diabetes is from south Asian countries such as India, Pakistan, and Bangladesh [1]. Individuals of SAS ancestry face a two-to-four-fold greater risk of developing type 2 diabetes, and its complications compared to Europeans (EUR), developing type 2 diabetes approximately a decade earlier and at comparatively lower BMI [3, 4]. This south Asian phenotype was also observed in studies of first-generational and multi-generational migrant adults [4]. Findings from a study conducted in the United Kingdom (UK) found that among populations younger than 30 years, the incidence of type 2 diabetes was almost three-times higher in SAS than in EUR participants [5]. By the age of 70 years 30-40% of SAS have type 2 diabetes in the UK – the prevalence at least twice than in EUR [5].

Studies in EUR populations have highlighted differences in cardiometabolic risk between males and females [6, 7]. There is a need to identify better clinical predictors of type 2 diabetes and especially early onset type 2 diabetes risk, not only between different ancestry groups and sexes, but also within SAS populations — given evidence of genetic heterogeneity among ancestry subgroups [8]. Early identification of high-risk populations and targeted preventive interventions are particularly important in SAS, as evidence shows they transition from high-risk states to type 2 diabetes more rapidly than EUR [5]. For example, the annual progression rate from impaired glucose tolerance to type 2 diabetes in SAS is 12–18%, compared to 5–11% in EUR [5]. Additionally, they may be at higher risk for other cardiovascular diseases even before a diagnosis of type 2 diabetes [9].

BMI is heavily dependent on the assumption of an average proportion of lean-to fat mass that is poorly applicable to many non-European ancestries. Studies [10, 11] and clinical guidelines [12] have increasingly identified lower BMI thresholds for SAS when classifying obesity. Added to this are newer findings that demonstrate lower proportion of lean mass in SAS [13], meaning that a measure of weight gain alone does not reflect the same risk in SAS as it does in EUR. This is largely due to underestimation of unfavourable adiposity in these populations [14, 15], while measures of central adiposity such as waist-to-height (WHTR) or waist-to-hip ratios (WHR) may serve as better predictors across ancestries and sexes [14, 16, 17]. This is also reflected in the weaker correlation between higher BMI and younger age of diagnosis in SAS compared to Europeans [18].

Genome-wide association studies (GWAS) have facilitated the development of polygenic scores (PGS), which estimate an individual’s genetic risk for various diseases and traits [19]. While GWAS are more representative of populations of European ancestry due to lack of large non-European genetic cohorts, there is substantial transferability of PGS across ancestry groups [20]. PGS enable the identification of populations with high genetic susceptibility for some conditions and could help to elucidate the contributions of common genetic variants in the aetiology of diseases. The advancement of these methods has led to a growing number of genetic risk scores tailored to different anthropometric and adiposity measures. Previous studies tried to utilise these methods to find genetic contributors of early onset type 2 diabetes among SAS populations [8, 21]. Since adiposity is one of the main risk factors for type 2 diabetes, it is important to understand the role of genetics for adiposity indicators in the early-onset type 2 diabetes across populations of different descents.

The influence of genetic risk for adiposity on clinically measured BMI and other anthropometric measures across different ancestries or sexes has not been studied [22]. If there is a significant impact of genetics for adiposity on type 2 diabetes onset, given that populations of non-European ancestries are at greater risk of type 2 diabetes and other cardiovascular conditions at lower BMI thresholds [10, 11, 23], and because current preventive strategies to identify high-risk populations are mainly target BMI, it is crucial to understand the extent to which clinically measured BMI explains or mediates this relationship. We hypothesise that measured BMI could be less effective in explaining the risk for early type 2 diabetes onset associated with genetic risk for BMI in individuals of SAS ancestry than in EUR, as they are more susceptible for central adiposity [24]. In such cases, relying solely on BMI may lead to an underestimation of their true risk. Polygenic scores offer an opportunity to build informed hypothesis on better anthropometric measures of risk with clinical impact.

Given the earlier onset of type 2 diabetes in SAS populations and the limitations of BMI in capturing adiposity-related risk, we explored whether genetic predisposition to adiposity, namely BMI, contributes to earlier disease onset in SAS and EUR ancestries. We examined how well measured BMI mediates this relationship across ancestries, and whether alternative adiposity-related polygenic scores offer better predictive value.

## Methods

### Genes & Health cohort

Genes & Health (G&H) is a community-based cohort of British Bangladeshi and British Pakistani individuals living in the UK, aged 16 years or above, recruited primarily in community and healthcare settings since 2015 [25]. At recruitment, each participant completes a brief questionnaire and consents to lifelong electronic health record (EHR) access, and to the use of their genetic data derived from donated saliva samples [25]. The recruitment process and detailed description of the cohort can be found elsewhere [25]. For this study, we used the genotype data of the January 2024 release on 41,631 participants who had both primary and secondary care data [20, 25]. Participants with type 2 diabetes and with complete data on BMI at diagnosis (measured within one-year before diagnosis and three months after) and available enhanced PGS for BMI were selected. The process for selecting the study population is shown in **Fig. S1**.

### Ethics

G&H operates under ethical approval (14/LO/1240), from London South East National Research Ethics Committee of the Health Research Authority (September 2014) [25].

### Genotype data, imputation, and quality control in Genes & Health

Genotyping was performed using the Illumina Global Screening Array v2 platform. Genetic ancestry was determined using principal component analysis (PCA). Imputation was done using a multi-ancestry TOPMED-r3 reference panel to genome build GRCh38. Variants with call rate <0.99 and minor allele frequency <1%, and single nucleotide polymorphisms with imputation quality score <0.3 were excluded. A detailed quality control process is described elsewhere [26]. Individuals who had no data on date of birth, sex and NHS number were removed (n=376).

### UK Biobank

UK Biobank (UKB) cohort was used to compare the mediating effect of BMI between enhanced BMI-PGS and the age at type 2 diabetes diagnosis in EUR populations. The recruitment, cohort profile of UKB population and genotyping was previously described elsewhere [27]. All pre-processing and analysis including eligibility criteria were the same as in G&H (**Fig. 1**). Analyses in the UKB were conducted under application ID 153692.

**Fig. 1.**
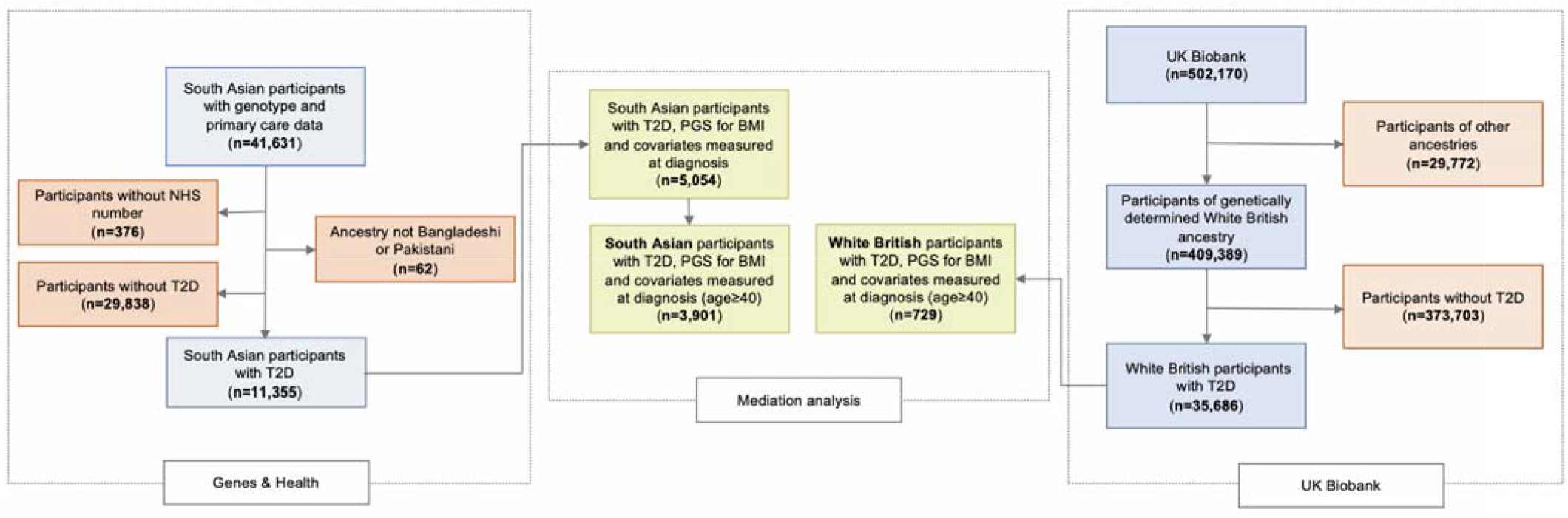
Included participants from the Genes & Health and the UK Biobank cohorts. This flow diagram illustrates the number of participants from the Genes & Health study and the UK Biobank included in each analysis. Since the minimum age of recruitment in UK Biobank was 40 years, for the mediation analysis, we restricted Genes & Health population with type 2 diabetes to those aged 40 or older. *Abbreviations: type 2 diabetes – type 2 diabetes; PGS – polygenic score; BMI – body mass index; NHS – National Health Service.

### Age at type 2 diabetes diagnosis

Type 2 diabetes was ascertained using SNOMED and ICD-10 codes extracted from participants’ EHRs (**Table S1)**. The codes used in this study were generated as a part of the NIHR AI Multiply consortium [28], and applied following previously established and published methodologies to define age at type 2 diabetes diagnosis [8, 26]. In the UKB, the age at type 2 diabetes diagnosis was determined using the same method as in the G&H cohort, with type 2 diabetes identified through pre-processed first occurrences files.

### PGS characteristics and calculation

For mediation analysis, we used enhanced BMI-PGS which was developed on external population and trained in the subsamples of UKB [29] and G&H. The reason behind using enhanced PGS is to overcome population overlap between development and evaluation cohorts, since most of the previously published PGS included individuals from the UKB for development. The methodology for generating PGS for UKB traits is documented elsewhere [29]. Enhanced BMI-PGS in the G&H was generated using the same algorithm as in UKB. In addition, we repeated the analysis for EUR participants using standard BMI-PGS [29], which was calculated for bigger population compared to the enhanced PGS but was not available for the G&H population. For validation of mediation analysis findings in the G&H population, additional best performing BMI-PGS was used (described below).

We also assessed the contribution of other adiposity PGS in age of type 2 diabetes diagnosis among SAS. We selected eight different PGS - for BMI, WC, WHR, hand grip strength (HGS), body fat (BF), trunk fat (TF), visceral adipose tissue (VAT) and gluteofemoral fat (GFAT) from the PGS catalogue website [30]. The selection process is described in **Text S1** and **Table S2-3**. Even though HGS is not considered as a direct adiposity measure, we included it because it indirectly conveys information about muscle strength, as previous studies suggest that SAS not only have higher visceral fat, but also lower muscle mass [13, 31]. All PGS, except for BMI, were fully developed on UKB population (n=7). Full characteristics of selected PGS are shown in **Table S4**. None of chosen PGS had overlapping populations with G&H. Using these PGS, we calculated individual genetic risk scores for British Bangladeshi and British Pakistani individuals using PLINK 2.0 software adjusting for age at recruitment, sex and 20 PCs. Their standardised measures (mean 0, SD 1) were utilised in this study.

#### Covariate definitions

We used routinely measured variables such as BMI (to capture obesity), total cholesterol (TC), HDL and alanine aminotransferase (ALT, to capture liver/visceral fat [14]) measured within one year before or three months after type 2 diabetes diagnosis were chosen for adjustment in the mediation analysis to reduce residual confounding. All other potential confounding variables had high missingness (>40%) in our sample and were therefore not used. TC and HDL were used to calculate non-HDL cholesterol. We restricted the selection of values measured up to three months after type 2 diabetes diagnosis to eliminate the possible effect of treatment and lifestyle changes which could follow diagnosis. We observed a non-differential selection bias for measurements of WC – participants with WC measurements were older, more likely to be males and had type 2 diabetes, p-value<0.001 (**Table S5**. Therefore, only BMI was used in mediation analysis. Complete case analysis was used instead of multiple imputation, as missingness in primary care data is unlikely to be completely at random. In such situations, complete case analysis can yield valid results, provided that the probability of missingness is not related to the outcome [32].

### Statistical analysis

#### Descriptive statistics

For descriptive statistics, the mean and standard deviation values for each continuous variable and counts with percentages, for categorical variables were estimated. The relationships between BMI at diagnosis and genetic risk for BMI (categorised as top decile vs. the rest) were compared using a t-test across ancestries. Linear regression was utilised to check associations between enhanced BMI-PGS with age at type 2 diabetes diagnosis and measured BMI.

#### Mediation analysis

We propose to use mediation analyses to estimate the direct and indirect effects of enhanced BMI-PGS on early onset type 2 diabetes. Indirect effect shows the proportion of total effect of genetic risk for BMI on earlier type 2 diabetes onset explained by changes in clinically measured BMI, while direct effect estimates the effect of BMI-PGS on age of type 2 diabetes via pathways other than clinically measured BMI. We examined to what extent genetic risk for BMI contributes to earlier onset of type 2 diabetes through changes in clinically measured BMI, in ancestry- and additionally sex-stratified mediation analysis using *process* package in R [33]. To reduce residual confounding and to control for population stratification, mediation analysis was adjusted for non-HDL cholesterol, ALT and 20 PCs in G&H and 10 PCs in UKB. 95% confidence intervals for indirect effect were estimated using bootstrapping method (n=500). Additionally, we examined the differences in the indirect effect between participants in the top decile for BMI-PGS versus remaining 90% for both SAS and EUR individuals. Where both total and indirect effects had the same direction of effect, we estimated the proportion of effect explained via mediator – indirect effect.

After this, to understand which PGS better differentiate extreme risk for BMI, WC or WHTR, we assessed the relationship between adiposity PGS and above-mentioned anthropometric measures using multivariable linear regression comparing populations at the top vs. bottom decile for PGSs.

#### Multivariable analysis

Since all other adiposity PGS were developed with UKB, we could not apply them for evaluation in the same cohort. Therefore, we restricted this analysis to G&H participants. To find which PGS are independent predictors of type 2 diabetes onset among SAS, ancestry- and sex-stratified multivariable linear regression models with all eight PGS adjusted for 20 PCs were built. To decide which PGS strongly contribute to the earlier onset of type 2 diabetes, we estimated partial R^2^ (variance explained by each variable in the model). To select the best combination of PGS for each subgroup, partial F test for nested models was estimated with backward selection approach. Since PGSs are constructed using common genetic variants which could potentially have pleiotropic effect, multicollinearity between included PGS was assessed by variance inflation factor (VIF). The goodness of fit for models was evaluated with Akaike Information Criterion (AIC). Ancestry- and sex-stratified relationships were checked. Following this, the associations for all significant PGS were compared between individuals in the top decile for genetic risk vs. the remaining 90% of population. The interaction test was conducted to check for ancestry differences. As the purpose of this analysis was to identify best genetic scores for adiposity measures predicting age of type 2 diabetes diagnosis and not developing a comprehensive prediction model, we did not assess models’ discriminative ability as genetic risk scores alone explain only small proportion in the trait variability.

#### Software

Python version 2.7.18 and R statistical software version 4.3.3 and PLINK 2.0 were used for data pre-processing and analysis, respectively.

### Reporting

This study is reported according to the STrengthening the REporting of Genetic Association Studies (STREGA) reporting recommendations [34].

## Results

### Study population and baseline characteristics

We included 5,054 G&H volunteers of British Pakistani and British Bangladeshi ancestry with type 2 diabetes to investigate the relationships between enhanced BMI-PGS and the age at type 2 diabetes diagnosis, and 729 UKB participants of EUR ancestry with type 2 diabetes who had data on enhanced BMI-PGS (**Fig. 1**). Since population in the UKB cohort is older than 40 years, the comparison in the mediation analysis was restricted to SAS participants aged 40 years or older (n=3,901). Median age at type 2 diabetes diagnosis was 50.5 years among SAS, and 64.0 years among EUR participants (p=0.95). These two populations did not have significant differences in covariates measured at diagnosis, except UKB population had significantly higher proportion of males than in G&H (p<0.001) (**Table S6**). Despite that, in each age group at type 2 diabetes diagnosis, SAS had lower median BMI than EUR (**Fig. S1**). Distributions of PGS used in this study are shown **Fig. S2-3**.

### Genetic risk for BMI is associated with earlier type 2 diabetes onset

One SD increment in the enhanced BMI-PGS was associated with earlier age at type 2 diabetes diagnosis and higher BMI at diagnosis in both ancestries (**Table S7a**). High clinically measured BMI was associated with earlier age at type 2 diabetes diagnosis in both SAS and EUR (**Table S7b**). To look at the effect of extreme genetic risk for BMI on BMI at type 2 diabetes diagnosis, we compared ancestry differences in BMI between individuals in the top decile of the enhanced BMI-PGS and the remaining population (**Fig. S4**). This difference in BMI was more pronounced among EUR, who had a four-point difference in median BMI between groups, compared to a one-point difference among SAS (**Fig. S4**).

### Mediation analysis: measured BMI fully explains genetic effect in white Europeans, only partially in south Asians

To assess how much of the estimated effect of the enhanced BMI-PGS on age at type 2 diabetes diagnosis was mediated by measured BMI we utilised ancestry- and sex-stratified mediation analysis method adjusting for non-HDL cholesterol, ALT and study-specific PCs. We estimated total, direct (not via BMI) and indirect (via BMI) effects. Indirect effect is the effect of BMI-PGS on age at type 2 diabetes onset explained by a change in individual’s BMI, while direct effect is the effect via other pathways. Total effect is overall effect of BMI-PGS on age at type 2 diabetes onset, and it is the sum of direct and indirect effects. While, per one SD increment in the BMI-PGS its total effect on age at type 2 diabetes diagnosis was −0.73 (95%CI −1.01; −0.45) among SAS and −0.57 (−1.05; −0.08) among EUR (**Fig. 2a-c** and **Table S8-9**), measured BMI explained only 28% of this association among SAS and the full effect among EUR (**Table S8-9**). After splitting the total effect to direct and indirect effects, the direct effect of BMI-PGS on age at type 2 diabetes onset became not significant in EUR subjects and indirect effect decreased to −0.33 (95%CI −0.49; −0.16), meaning that the effect of genetic risk for BMI affects earlier type 2 diabetes onset only via changing BMI in this group. Results of unadjusted analysis were analogous (**Table S10**). Similar findings were observed when standard BMI-PGS was utilised in EUR (**Table S8**) and additional BMI-PGS (PGS000027) in SAS populations (**Table S11**). Indirect effect was less strong among whole population with type 2 diabetes in SAS (without age restriction) (**Table S12**). In addition, the strength of indirect effect was stronger among participants in the top decile compared to the remaining populations in both ancestries: −0.42 (95%CI −0.62; −0.26) among SAS and −0.78 (95%CI −1.35; −0.34) among EUR (**Table S13**). In the sex-stratified analysis, despite similar total effects, the indirect effect of BMI-PGS on age at type 2 diabetes onset via BMI was stronger among females than in males in both ancestries, though the effect among SAS males became not significant (**Tables S8-9**).

**Fig. 2a-d.**
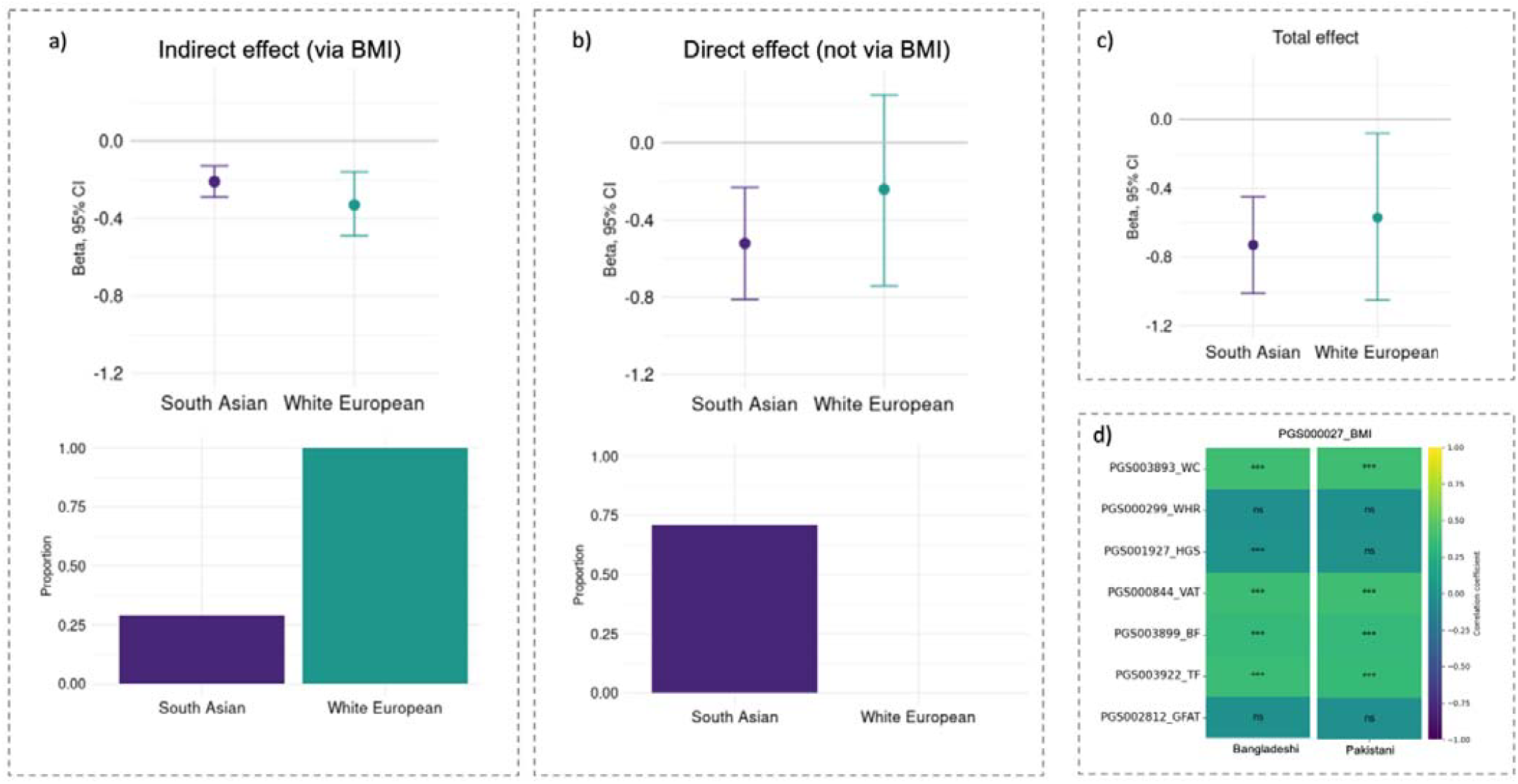
Mediation analysis results show that BMI-PGS influences earlier age at type 2 diabetes onset fully via increasing BMI in white European participants, and only partial y via this path in south Asians. **(a, b, c)** Ancestry-stratified results of mediation analysis and **(d)** correlation between BMI-PGS and PGSs for different adiposity measures. **(a, b, c)** Upper plots show beta-coefficients with 95% CI for indirect (BMI), direct (not via BMI), and total effects per SD increment in BMI-PGS on age at type 2 diabetes diagnosis for south Asian (n=3,901) and white European participants (n=729). **(a, b)** Bar plots below illustrate the partitioned proportion (in %) of total effect into indirect and direct effects by ancestries. **(d)** Heatmap shows correlation between BMI-PGS with other PGSs for adiposity measures among south Asians. Strongest correlations were observed for WC, VAT, Trunk Fat and Body Fat. Indirect effect: effect of BMI-PGS on age at type 2 diabetes onset via increasing BMI; direct effect: the effect of BMI-PGS on age at type 2 diabetes not via increasing BMI, which could be via other pathways or natural direct effect.

### BMI-PGS is moderately correlated with central adiposity scores

The observed association between BMI-PGS and age at type 2 diabetes diagnosis (total effect) was slightly stronger among SAS than in EUR, but since confidence intervals overlapped difference was not statistically significant (**Table S8-9**). This finding suggests that for SAS, a bigger proportion of the effect of genetic risk for BMI on type 2 diabetes diagnosis is not fully explained by changes in measured BMI. It is likely this effect is driven by variants in the BMI-PGS that represent true signals for adiposity. In SAS, correlation analysis between BMI-PGS with other PGS for adiposity indicators demonstrated that BMI-PGS is moderately correlated with PGSs for WC, BF, TF and VAT (**Fig. 2d**). This could potentially be a partial explanation for the effect of BMI-PGS on type 2 diabetes diagnosis in this ancestry group. Due to limited availability of WC measurements and likely selection bias related to who this was measured in, we could not test these pathways directly in our cohort.

### BMI-PGS poorly predicts central adiposity compared to other PGSs

In order to explain the strongest polygenic predictors for measured BMI, WC and WHTR, we compared these measures between populations in the top vs. bottom deciles for adiposity PGSs among SAS. BMI-PGS could not discriminate higher central adiposity measures such as WC and WHTR among SAS individuals with type 2 diabetes. Despite poor recording of central adiposity indicators (**Table S14**), we observed that WC-PGS, TF-PGS and BF-PGS, but not BMI-PGS, were better at discriminating clinically measured WC, WHTR and BMI (**Fig. 3, Table S15**).

**Fig. 3a-c.**
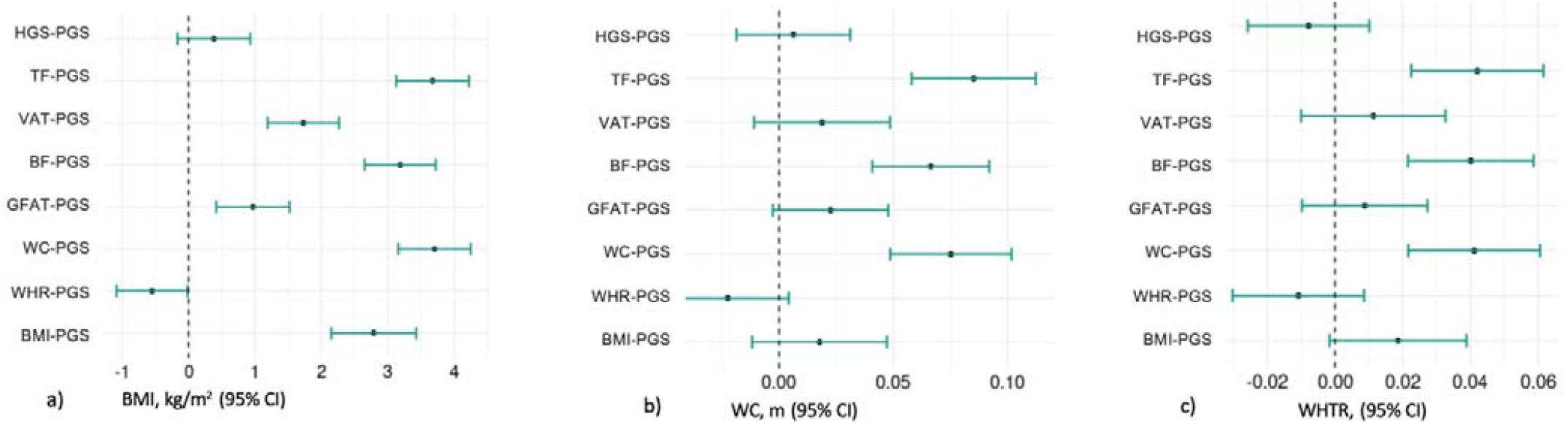
Comparison of BMI, WC and WHTR at genetic extremes for adiposity measures. **a)** Difference in BMI in kg/m^2^ between individuals in the top decile vs. bottom decile for adiposity PGSs (n=505 in each decile). **b)** Difference in WC, in m, between individuals in the top decile vs. bottom decile for adiposity PGSs (n=135 in each decile). **c)** Difference in WHTR between individuals in the top decile vs. bottom decile for adiposity PGSs (n=128 in each decile). These forest plots indicate the limited ability of BMI in discriminating central adiposity measures among south Asian populations. *Abbreviations: HGS – hand grip strength, TF – trunk fat, VAT – visceral adipose tissue, BF – body fat, GFAT – gluteofemoral fat, WC – waist circumference, WHR – waist-to-hip ratio, BMI – body mass index, WHTR - waist-to-height ratio

### Different adiposity PGS explain greater variance in type 2 diabetes onset in south Asian sub-populations

We performed ancestry- and sex-stratified models adjusted for multiple PGS for adiposity indicators. Based on the results of backward selection approach, in British Pakistanis along with BMI-PGS, WHR-PGS was a significant predictor of earlier age of type 2 diabetes onset in males and females (**Fig. 4a-c**). In contrast, in British Bangladeshis, in addition to BMI-PGS, WC-PGS among males and GFAT-PGS and BF-PGS among females were better predictors of age of type 2 diabetes onset among individuals of British Bangladeshi descent (**Fig. 4a-c**). These models showed the lowest AIC statistics indicating better goodness of fit and all PGSs in models had VIF<2 indicating the absence of multicollinearity. In all subgroups BMI-PGS explained the highest variance in the age at type 2 diabetes diagnosis based on partial R^2^ values, followed by PGS for central adiposity measures such as WC among Bangladeshi participants and WHR among Pakistani individuals (**Fig. 4a-c**).

**Fig. 4a-c.**
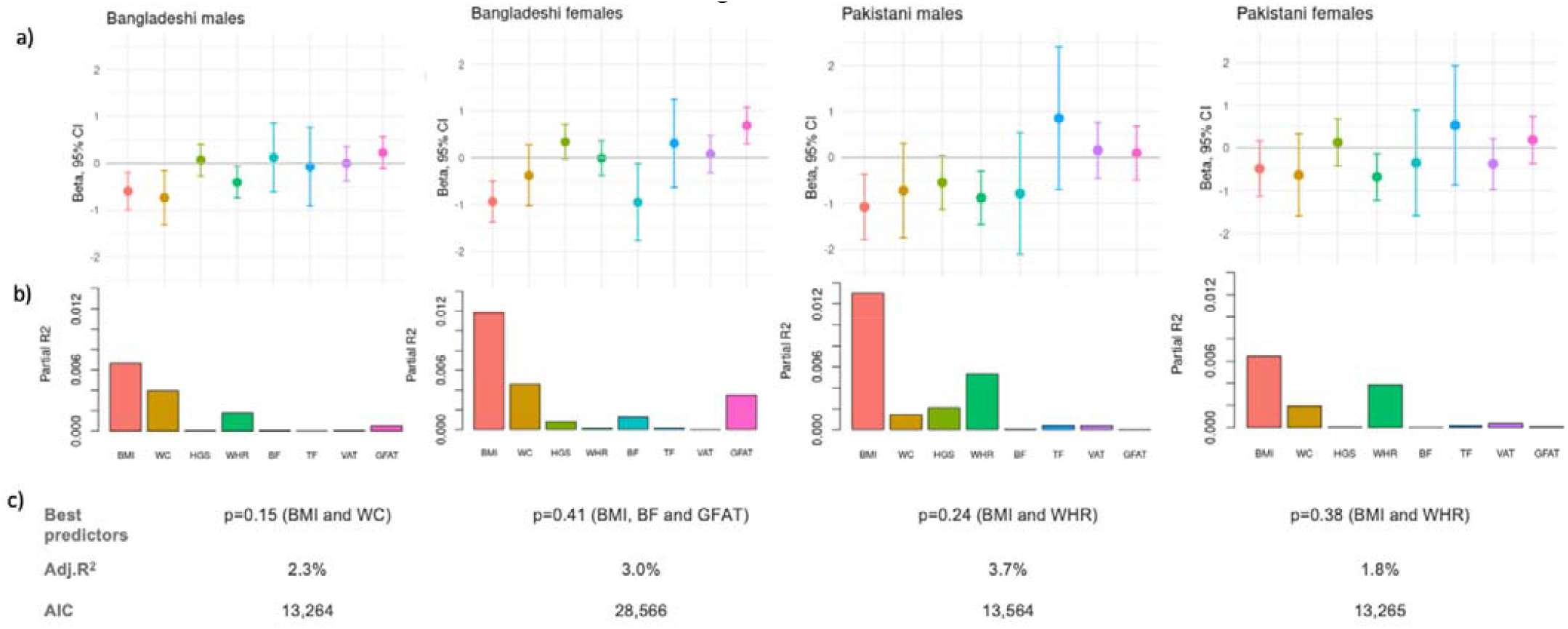
Best PGSs predicting age at type 2 diabetes diagnosis across ancestries and sexes varies. **(a)** Ancestry- and sex-stratified associations between PGSs for anthropometric and adiposity measures and age at type 2 diabetes diagnosis (adjusted for 20 PCs). **(b)** Bar plots show variance explained in the age at type 2 diabetes diagnosis by each PGS in these stratified models. **(c)** A table below shows the best combination of PGSs predicting the age at type 2 diabetes onset across ancestries and sexes based on partial F-test, AIC and adjusted R2. PGS for BMI and WHR were best predictors of age at type 2 diabetes onset among participants of British Pakistani background. Among British Bangladeshi individuals we observed sex differences – BMI-PGS and WC-PGS among males and BMI-PGS, BF-PGS and GFAT-PGS among females. *Abbreviations: BMI – body mass index; WC – waist circumference; BF – body fat; GFAT – gluteofemoral fat; WHR – waist-to-hip ratio.

### Genetic risk for central adiposity is a stronger predictor in British Pakistanis than British Bangladeshis

In the sex-stratified analysis, we observed significant interaction between ancestry and WHR-PGS in relation to the age at type 2 diabetes onset (p=0.03 for interaction test), where the effect was significantly stronger among British Pakistani males and females compared to individuals of British Bangladeshi background - ß= −0.92 (p<0.001) vs. ß= −0.47 (p=0.004) for males and ß=-0.72 (p=0.008) and ß=-0.13 (p=0.48) for females (**Fig. S5, Table S16**). The protective effect of GFAT-PGS was observed only among British Bangladeshi females with ß=0.61, p<0.001. The increment in BMI-PGS, WC-PGS and BF-PGS was associated with earlier type 2 diabetes diagnosis in all subgroups (**Fig. S5, Table S16**). As these PGSs were less predictive of central adiposity measures compared to their ability to predict BMI in our SAS population (**Table S2-3**), we expected low discriminative ability for central adiposity and therefore did not utilise them for mediation analysis.

## Discussion

### Summary of key findings

This study included British Bangladeshi and British Pakistani individuals from the Genes & Health cohort and white Europeans from the UK Biobank cohort and tried to examine the relationships between genetic risk for adiposity and early onset type 2 diabetes, as well as to understand what proportion of that effect is mediated via clinically measured BMI. Our findings showed that high genetic risk for raised BMI could have a significant impact on earlier onset of type 2 diabetes among both SAS and EUR populations. While the total effect of BMI-PGS on age of diagnosis was explained by increase in recorded BMI in EUR, BMI explained only 28% of the effect of BMI-PGS among SAS subjects, leaving a bigger proportion unexplained. In both ancestries, the effect of BMI-PGS on early onset type 2 diabetes which was mediated by measured BMI was significantly stronger among populations in the top decile compared to the remaining population and among females than in males. The polygenic basis of adiposity was better explained by combination of WC and TF in SAS. We therefore extended our analyses to demonstrate that along with BMI-PGS other PGS for central adiposity measures could be important contributors in the development of type 2 diabetes at younger ages among SAS ancestries. Lastly, this study demonstrated that the mechanisms of effect of different genetic scores for adiposity indicators could differ between British Pakistani and British Bangladeshi populations. Ancestry significantly moderated the effect of genetic risk for WHR on age of type 2 diabetes diagnosis, emphasising a significantly stronger role of genetic risk for central adiposity on age of onset among British Pakistanis than among British Bangladeshis.

We used mediation analysis to understand if clinically measured BMI adequately captures the genetic risk for BMI and early onset type 2 diabetes, and if there were differences in this relationship across ancestry groups (SAS and EUR). These findings have implications for the validity of clinically measured BMI as a risk predictor for early-onset type 2 diabetes. Our findings suggest the effect of BMI-PGS among SAS could predominantly drive type 2 diabetes risk through central adiposity measures rather than BMI. Measures of central adiposity are poorly captured in routine clinical care and were unavailable in our cohort, underscoring the need for future studies to explore their role more fully. Significant moderate correlations between BMI-PGS and PGS for central adiposity measures in SAS might partially support this inference.

Previous observational studies [16, 17, 35], as well as the latest recommendation for weight management of the UK National Institute for Health and Care Excellence (NICE) which was published in 2023 [36] advised healthcare practitioners to use WHTR alongside BMI to assess risk for cardiometabolic diseases, especially among populations of diverse ancestries. This is justified by the superior ability of WHTR to capture changes in the visceral area, since height is relatively stable during adult life, as well as similar predictive ability is across sexes. This is important because compared to EUR populations, SAS are more prone to have unfavourable adipose distribution such as a preponderance of visceral and ectopic fat, which is not well captured by BMI and which is associated with dyslipidaemia, hypertension, insulin resistance and lower amounts of gluteofemoral fat [37, 38]. Neel et al. suggested the ‘thrifty genes’ hypothesis suggesting that historical exposure to famine may have le certain populations to develop a genetic predisposition fat storage, particularly in the visceral area [39]. Therefore, high genetic risk for BMI in these populations potentially could lead not only increased overall adiposity but also to a greater accumulation of central or ectopic fat.

### Sex- and ancestry-specific findings

Sex differences in the effect of genetic risk for BMI on type 2 diabetes onset via clinically measured BMI were observed in both SAS and EUR. Despite similar total effects, indirect effect was stronger in females than in males (**Table S7-8**). This potentially could be explained by protective effect of oestrogens among pre-menopausal females, among whom fat tend to be peripheralised, accumulating in subcutaneous adipose tissues compared to visceral and ectopic deposition among males [40, 41]. This could be associated with an increase in overall adiposity – BMI – among females, and central adiposity indicators among males. Remaining direct effect among both males and females of SAS background but not in EUR, might indicate higher significance of central adiposity pathways or other biological mechanisms in this population which are not well captured by BMI. Previous study using partitioned PGS (pPGS) showed that along with genetic predisposition to earlier beta-cell dysfunction, pPGS for unfavourable fat distribution, namely high genetic risk for lipodystrophy 1 (insulin resistance) and lipodystrophy 2 (ectopic fat, liver fat) clusters, were significant drivers of early-onset type 2 diabetes among SAS populations [8]. Taking into account these findings, it is possible that the effect of BMI-PGS on early onset and therefore diagnosis of type 2 diabetes in our study could be due to several reasons – effect on BMI, effect on ectopic fat accumulation, which may or may not be reflected in central adiposity measures, other mechanisms and potential pleiotropic effect due to population stratification. Taken together with the fact that all PGS were developed predominantly in EUR populations limits us from conducting a Mendelian randomization study.

Our findings suggest that the impact of overall and central adiposity among individuals of Bangladeshi and Pakistani descent, highlighting potential variation in the genetic architecture of obesity within SAS populations. Larger, better powered studies are needed to further understand the genetic architecture of unfavourable adipose distribution in SAS ancestries. It is essential to examine how genetic risks for adiposity indicators influence changes in overall and central adiposity across different ancestries over the life course and in different environment. This includes, comparing migrant and non-migrant communities, as environmental factors may modulate the relationship between PGS and anthropometric measures [42]. The two-to-four-year difference in type 2 diabetes onset between individuals in the top decile for genetic risk of various adiposity indicators and the rest of the population suggests they may benefit from earlier lifestyle interventions. Furthermore, as previous research has shown that the effectiveness of glucose-lowering medications in SAS may vary by genetic risk for type 2 diabetes [8], it is important for future studies to investigate whether their effectiveness also differs by genetic risk for various adiposity indicators, and whether certain drug classes may be more effective in managing the disease. In our cohort, almost 80% of SAS with type 2 diabetes in both top and bottom deciles of BMI-PGS were prescribed metformin (**Fig. S6**). In addition, a growing body of evidence suggests a significant inverse association between BMI-PGS and the effectiveness of weight loss interventions, where weight loss is assessed using BMI [43, 44]. Our findings underscore the importance of considering alternative measures of metabolic health — such as WHTR or other direct indicators of central adiposity — along with BMI when evaluating the effectiveness of these interventions in diverse populations [22].

Considering the low representativeness of the diverse world populations in genetic discovery studies and resulting PGS, the true effect of adiposity PGSs on age at type 2 diabetes onset among subjects SAS descent might differ. All PGS used in our study, except for BMI, were developed utilising the UKB which predominantly includes participants of EUR background. This lopsided nature of available data emphases the significance of increasing the number of genetic studies involving diverse populations, including SAS. Increasing their representation will enhance the accuracy and effectiveness of genetic risk prediction tools in diverse populations, helping to ensure equitable benefit from scientific advances.

### Strengths and limitations

Some limitations should be considered when interpreting these findings. First, although the effect of BMI-PGS was assessed longitudinally, the relationship between BMI and age at type 2 diabetes diagnosis in the mediation analysis was assessed cross-sectionally. This limits our ability to infer temporality or causality in that pathway. Second, sample size of British Bangladeshi participants was almost two times bigger than of British Pakistani individuals which may have influenced the power to detect associations in subgroup analyses. Third, these results should be interpreted with caution, since G&H participants are not genetically representative of all SAS populations but still provide insight into the genetic interplay of adiposity-related type 2 diabetes risk in British South Asians. Fourth, due to limited sample size of SAS in UKB we were unable to validate our findings in an independent, ancestry-matched dataset of comparable size. Fifth, we utilised the date of type 2 diabetes diagnosis, which is not the actual date of disease onset, which could have happened earlier. However, in this case, this would only introduce bias towards the null and indicate a stronger relationship between genetic risk for adiposity and true age of type 2 diabetes onset. Finally, even though we used comparable enhanced BMI-PGS in mediation analysis, for other adiposity indicators we utilised PGS which were developed on populations with small proportion of populations from non-European ancestry backgrounds. Therefore, true effects could differ in SAS and future studies need to validate this finding in independent cohorts.

This study has several strengths. First, we utilised large cohort of SAS participants who are underrepresented in research and filled in the gaps in evidence related to genetic basis of obesity driven metabolic disease. Second, utilising an enhanced BMI-PGS allowed us to compare the genetic pathway across two ancestries. These findings were consistent when alternative PGS were applied among both ancestry groups. Third, the linkage of genetic data to participants’ longitudinal EHR allowed us to find differences in the associations between PGS for adiposity indicators and age at type 2 diabetes diagnosis between Bangladeshi and Pakistani populations.

### Conclusion and future perspectives

This study shows that both British Bangladeshi and British Pakistani and white Europeans with high genetic risk for BMI are at increased risk of early type 2 diabetes onset. However, while recorded BMI could largely explain the association in EUR it only could partially explain this impact among SAS. In SAS, WC-PGS, TF-PGS and BF-PGS provided better discrimination of overall and central anthropometric measures than BMI alone. Given the limited representation of SAS populations in the existing PGS development, the true effect of PGS on the age of type 2 diabetes onset among these populations could be underestimated. These findings suggest that adiposity-related genetic risk for early type 2 diabetes onset differs not only across different ancestries as well as between SAS subgroups. Earlier preventive lifestyle interventions targeting individuals with high genetic risk for adiposity could be beneficial in delaying type 2 diabetes onset in these high-risk populations.

## Data Availability

All data produced in the present study are available upon reasonable request to the authors

## Abbreviations

PGS: polygenic score
SAS: South Asian
EUR: White European
WC: waist circumference
WHTR: waist-to-height ratio
WHR: waist-to-hip ratio
VAT: visceral adipose tissue
BF: body fat
TF: trunk fat
GFAT: gluteofemoral fat
HGS: hand-grip strength
GWAS: Genome-wide association studies
G&H: Genes & Health cohort
UKB: UK Biobank
EHR: Electronic Health Records
NIHR: National Institute for Health and Care Research
TC: total cholesterol
ALT: alanine aminotransferase
PC: principal component
NICE: UK National Institute for Health and Care Excellence
pPGS: partitioned polygenic score

## Acknowledgements

We thank Social Action for Health, Centre of The Cell, members of our Community Advisory Group and staff who recruited and collected data from volunteers. We thank the NIHR National Biosample Centre (UK Biocentre), the Social Genetic & Developmental Psychiatry Centre (King’s College London), the Wellcome Sanger Institute and the Broad Institute for sample processing, genotyping, sequencing and variant annotation. We also thank Barts Health NHS Trust, NHS Clinical Commissioning Groups (City and Hackney, Waltham Forest, Tower Hamlets, Newham, Redbridge, Havering, Barking and Dagenham), East London NHS Foundation Trust, Bradford Teaching Hospitals NHS Foundation Trust, Public Health England (especially D. Wyllie), Discovery Data Service/Endeavour Health Charitable Trust (especially D. Stables), Voror Health Technologies (especially S. Don), NHS England (for what was NHS Digital) for GDPR-compliant data sharing backed by individual written informed consent. Most of all, we thank all of the volunteers participating in Genes & Health.

## Funding

B.O. is funded by the Wellcome Trust PhD programme - health data in practice: human-centred science (218584/Z/19/Z). S.H. is funded by Wellcome HARP Doctoral Fellowship 227532/Z/23/Z. R.M. and M.K.S. are funded by Barts Charity (MGU0504). S.F. is funded by the Tackling Multimorbidity at Scale Strategic Priorities Fund program (grant number MR/W014416/1) delivered by the Medical Research Council and the National Institute for Health Research (NIHR) in partnership with the Economic and Social Research Council and in collaboration with the Engineering and Physical Sciences Research Council. Analyses in the UK Biobank were conducted under application ID 153692. The funders of this study had no role in study design, data collection, data analysis, data interpretation or writing of the report.

## Conflict of interest

We declare no conflict of interest.

## Author contributions

B.O. conceived the idea of the study, extracted genetic and clinical data, analysed data, wrote first draft, reviewed and edited the manuscript. M.K.S., R.M. and S.F. supervised and contributed to designing methodology, interpretation of findings, reviewed and edited the manuscript. S.H., M.B. M.S. and J.Z. contributed to genetic data extraction, interpreted findings and edited the manuscript. All authors gave final approval of the final draft and have contributed to the manuscript.

## Data availability

Genes & Health: Researchers and industry partners globally can access individual-level participant data by submitting an application to the Genes & Health Executive (https://www.genesandhealth.org/), which reviews requests on a monthly basis. UK Biobank: Individual-level data are accessible to researchers through an application process via the UK Biobank (https://www.ukbiobank.ac.uk/).

## Guarantor Data Access and Responsibility Statement

B.O. is the guarantor of this work, had full access to all the data, and takes full responsibility for the integrity of data and the accuracy of data analysis.

## Notes

### Competing Interest Statement

The authors have declared no competing interest.

### Funding Statement

This study was funded by Wellcome Trust PhD programme - health data in practice: human-centred science (218584/Z/19/Z)

### Author Declarations

G&H operates under ethical approval (14/LO/1240), from London South East National Research Ethics Committee of the Health Research Authority (September 2014)

